# State tanning bed availability is associated with early-onset Melanoma incidence in the Midwest and Southern United States

**DOI:** 10.64898/2026.08.21.26361039

**Authors:** Danielle Graffam, Jason Semprini

## Abstract

Despite known carcinogenic properties, indoor tanning remains popular among young adults and may contribute to early-onset melanoma. Our study aims to compare early-onset melanoma incidence by state availability of tanning beds. We analyzed population-based melanoma incidence data (2019-2023) from the National Program of Cancer Registries and calculated Incidence Rate Ratios (IRR) using verified state-level quintiles of tanning bed availability. Overall, in the Midwest/South regions, melanoma incidence increased with greater tanning-bed availability, from 8.7 cases per 100,000 population in Quintile 1 to 14.8 cases per 100,000 population in Quintile 5 (IRR = 1.69; CI = 1.65–1.74). No such relationship was found in the Northeast/West regions. In conclusion, we found that in Southern and Midwest states, increased availability of tanning beds was associated with higher early-onset melanoma in non-Hispanic White males and females, in both metro and non-metro counties. Policies which reduce tanning bed availability in high utilization regions may have potential to reduce early-onset melanoma.

## Introduction

Between 1975-2005, the incidence of early-onset melanoma doubled, with rates persisting through 2023 (Supplemental Table 1). Despite known carcinogenic properties, indoor tanning remains popular among young adults and may contribute to early-onset melanoma^1,2^. Indoor tanning is prevalent among females (18%) and males (6%), particularly in the Midwest and Southern regions^3^. While some states have adopted policies restricting tanning bed use among youth, tanning bed availability appears to be a driving factor for tanning bed utilization among young people more generally^4^. Our study aims to compare early-onset melanoma incidence by state availability of tanning beds.

We analyzed population-based melanoma incidence data (2019-2023) from the National Program of Cancer Registries (https://www.cdc.gov/national-program-cancer-registries/index.html). Melanoma cases were restricted to non-Hispanic White adults, aged 20-49 in the South and Midwest regions. We then categorized states into quintiles (Supplemental Table 2) according to verified data on the number of indoor tanning beds per capita (Wehner). To compare early-onset melanoma incidence by state quintile group, we calculated incidence rate ratios (IRRs) using the lowest state quintile as the reference group. Each IRR calculation was stratified by sex and metropolitan status, with 95% confidence intervals (CI) based on Tiwari correction method.

Overall, between 2019-2023, 45,193 non-Hispanic White adults were diagnosed with early-onset melanoma (Table 1). The lowest overall quintile reported a rate of 8.7 cases per 100,000 population. Quintiles 4 and 5 reported a rate of 14.9 and 14.8 cases per 100,000 population, respectively. The lowest incidence rate was found among non-metro males in quintile 1 (6.6). The highest incidence rate was found among non-metro females in quintile 5 (20.5.).

**Table 1.**
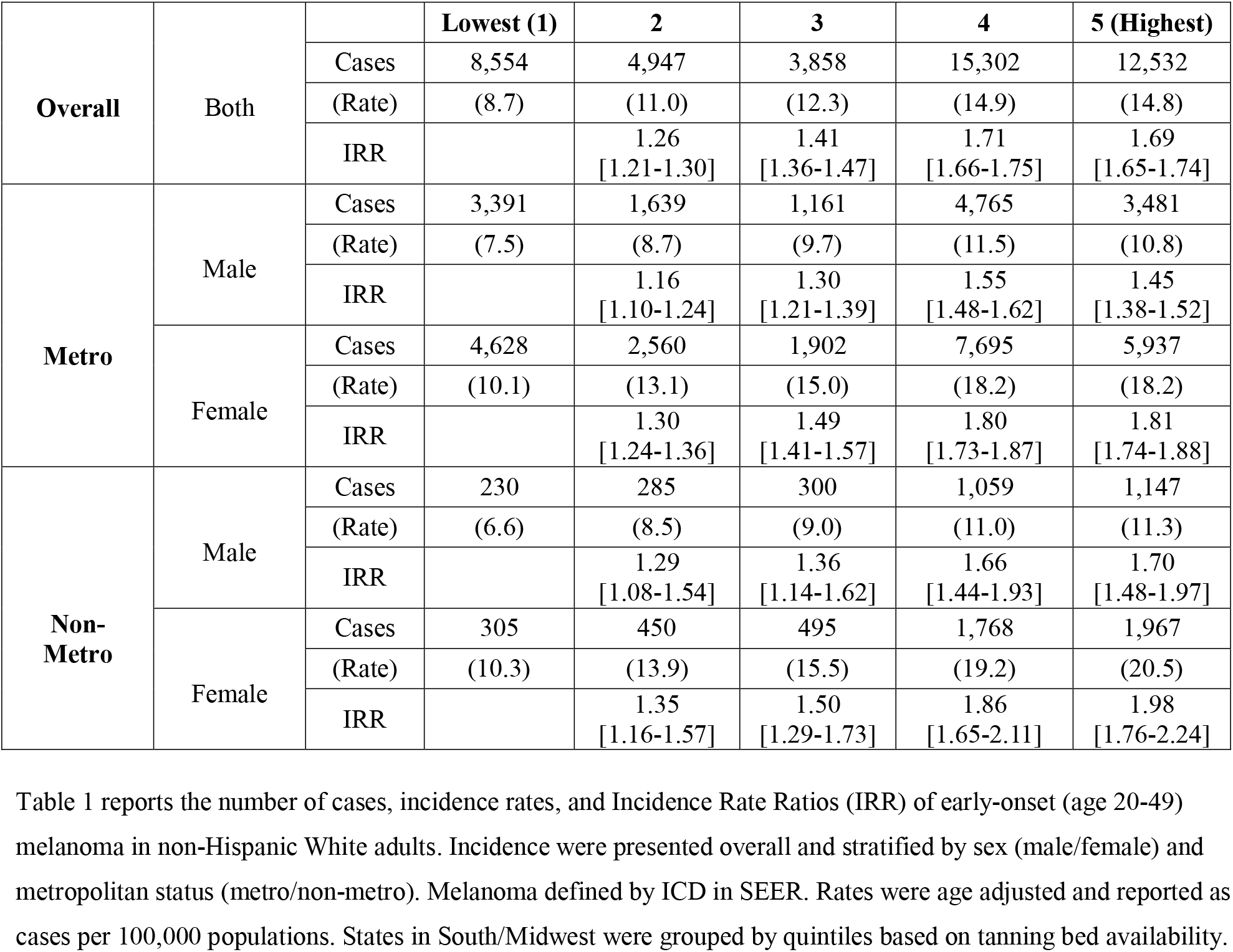
Early-Onset Melanoma Among Non-Hispanic White Adults (Age 20-49) in Southern and Midwest States by Sex and Metropolitan Status.

All IRR calculations were statistically significant, suggesting that state-level tanning bed availability was associated with higher incidence of early-onset melanoma (Figure 1). Overall, melanoma incidence increased with greater tanning-bed availability, from 8.7 cases per 100,000 population in Quintile 1 to 14.8 cases per 100,000 population in Quintile 5 (IRR = 1.69; CI = 1.65–1.74). The Quintile 5 associations within non-metropolitan counties were similar between males (IRR = 1.70; CI = 1.48-1.97] and females (IRR = 1.98; CI = 1.76-2.24). Within metropolitan counties, Quintile 5 associations were stronger in females (Male IRR = 1.81; CI = 1.74–1.88 and Female IRR = 1.98; CI = 1.76-2.24).

**Figure 1:**
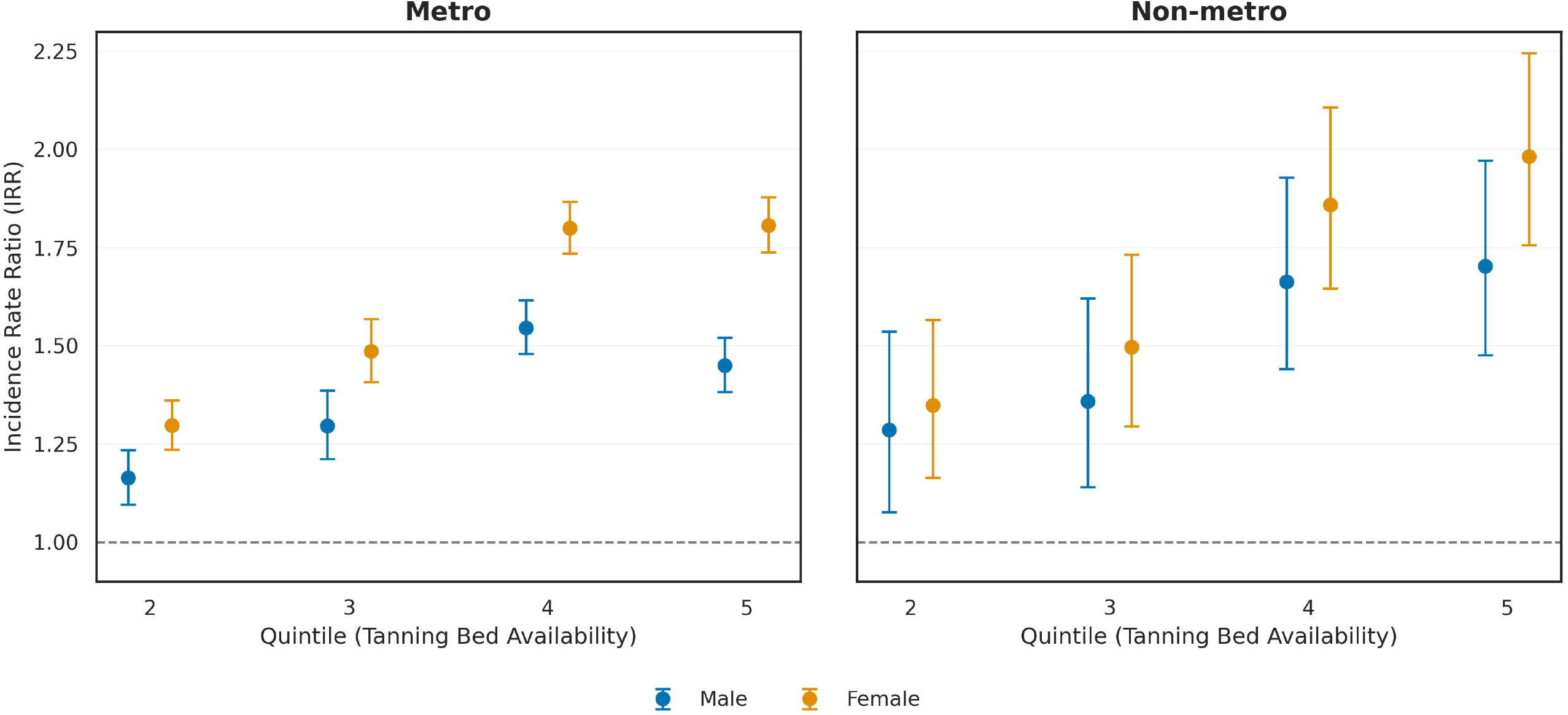
Incidence Rate Ratios (IRR) Comparing Early-Onset Melanoma Incidence by State Quintiles of Tanning Bed Availability. Figure 1 visualizes the association between state tanning bed availability and early-onset melanoma incidence. Quintiles represent state-level groupings of tanning bed availability per capita, with Quintile 1 representing the lowest availability and serving as the reference category. The study population was restricted to non-Hispanic White (NHW) melanoma cases aged 20–49 years residing in the Midwest or South census regions. Incidence rate ratios (IRRs) compare age-adjusted melanoma incidence rates in Quintiles 2–5 with Quintile 1; error bars represent 95% confidence intervals. Source: Surveillance, Epidemiology, and End Results (SEER) Program, 2019–2023.

We found that in Southern and Midwest states, increased availability of tanning beds was associated with higher early-onset melanoma in non-Hispanic White males and females, in both metro and non-metro counties. A limitation of our study relates to its ecological design, which prohibits evaluating the link between tanning bed use and incident cases of melanoma. State-level differences in melanoma risk may also be confounded by other factors and our observed associations do not generalize to lower tanning bed utilization regions (Supplemental Table 3). Still, despite these limitations, by merging nationally representative, population-based cancer data with verified tanning bed availability data, we advance the evidence base suggesting that greater tanning bed availability may increase early-onset melanoma^5^. Therefore, policies which reduce tanning bed availability in high utilization regions may have potential to reduce early-onset melanoma.

## Data Availability

Data is publicly available from NPCR.

https://www.cdc.gov/national-program-cancer-registries/index.html

